# A COVID-19 epidemic model integrating direct and fomite transmission as well as household structure

**DOI:** 10.1101/2020.04.25.20079178

**Authors:** Karunia Putra Wijaya, Naleen Ganegoda, Yashika Jayathunga, Thomas Götz, Wolfgang Bock, Moritz Schäfer, Peter Heidrich

## Abstract

This paper stresses its base contribution on a new SIR-type model for COVID-19 including direct and fomite transmission as well as the effect of distinct household structures. To what extent increasing the physical-distancing-related contact radius and enhancing mass control (public curfew, lockdown, workplace clearance, and school closure) reduce the number of predicted active cases is studied via parameter estimation.

## 1. Introduction

In December 2019, a number of residents were diagnosed with pneumonia in Wuhan, China alarming a viral outbreak. Later in early January 2020, it was identified that the cause of this pneumonia be a novel coronavirus (SARS-CoV-2) and the disease is officially named as COVID-19 (Huang et al., 2020). World Health Organization (WHO) declared COVID-19 as a pandemic on March 11, 2020 and has been conveying technical guidance to mitigate the disease burden with over 20 million confirmed cases and over 125,000 fatalities worldwide as of mid April, 2020 (World Health Organization, 2020c). Quantification measures such as the basic reproductive number and case fatality rate direct scientists, authorities, and general public toward testing and simulating, bearing interventions and evaluating healthcare capacity, and preventive habits, respectively.

Signs and symptoms of the infection start from dry cough and fever, and then some other mild clinical manifestations would be possible until severe levels such as acute respiratory distress syndrome persist (Chen et al., 2020). According to early surveillance, Huanan seafood market was the originating place and thereafter human-to-human transmission has escalated in an exponential pace. As per RNA-sequence analysis, the novel coronavirus may be arisen from cross-species transmission between bats and snakes (Ji et al., 2020). Three variants of the virus were identified in a phylogenetic network analysis that used bat virus as an outgroup (Forstera et al., 2020). Two of them are common in Europeans and Americans, while the other one is common in East Asia, showing the vulnerability in genetic changes. Li et al. (2020a) as per their analysis of early dynamics, reported that the basic reproductive number ℛ_0_ of COVID-19 in China was 2.2 and incubation period 5.2 days. In another early estimation, Zhao et al. (2020) reported that ℛ_0_ ranges from 2.24 to 3.58. Amidst those varying estimations, transmission risk of COVID-19 is noteworthy as compared to previous two outbreaks SARS in 2003 and MERS in 2012 since individuals can be infectious before being symptomatic (Huang et al., 2020; Hellewell et al., 2020).

After the early signs of the outbreak, health authorities worldwide have taken preventive actions with the patronage of WHO (World Health Organization, 2020a). These actions range over good practices in household level such as washing hands, wearing masks and undertaking social distancing in public level, quarantine at home or designated centers, travel restrictions in international level, and finally lockdowns. Countrywise variations can be observed in motivation, compliance, and success toward effective control strategies (United Nations, 2020). There is notable difference in hospitalization, for instance China practiced it as an isolation procedure as well, while USA and UK recommended it only for severe cases (Verity et al., 2020). Whether to extend or relax distancing measures is still a dilemma as there is an emerging threat of second wave resurgence (Leung et al., 2020).

Four levels of transmission designated by WHO (1–no cases, 2–sporadic cases, 3–clusters of cases, 4–community spread) set the need of basic interventions and intensity (World Health Organization, 2020b). Countries are given a more detailed classification, indicating lower to higher risk as interrupted transmission, under investigation, imported cases only, local transmission and community transmission. However, it should be scaled down to workplace and household level. Then compartmental aspect of how many are susceptible, symptomatic, asymptomatic, recovered, etc. will decide the immediate risk to the community (Chan et al., 2020). Both susceptibility and fatality risks have age dependency, where older group shows higher vulnerability (Wu et al., 2020; Zhou et al., 2020). In addition, sneezing, touching contaminated surfaces, individual immunity, number of members sharing the space necessarily come into effect. To be more successful, all preventive measures should be communicated in a practically sounding way, irrespective of the socio-economic background of community or country. Under dreadful circumstances, we should focus on what are expected from each individual to mitigate further risk of COVID-19.

In this work, we are concerned with a certain rule of thumb that can be applied on individual level. Two measures are put under investigation: minimum contact radius during physical distancing and the number of cross-household encounters. The first measure has been appearing in the media from various countries, steering the possibility of getting infection via direct transmission (i.e. by sneezing). The second measure represents how many encounters with infected persons per day a susceptible person makes on average. This number is unobservable, yet is known to drive the force of infection in standard epidemic models, therefore also subject to treatment. After model derivation and data fitting, our final plan will be to compare the performance of extending the minimum contact radius and reducing the number of cross-household encounters from the model solution in terms of minimizing the number of predicted active cases. Our model bases its design on a standard SIR-type model with additional features highlighting minimum contact radius, direct and fomite transmission, also different household structures incorporating district-specific attributes. Via the latter we can be more specific on selecting transmission routines within and between households. Many factors are infused into a basic model through parameters in the form of rates, probabilities, and ratios, as seen shortly.

## 2. Model description and accompanying assumptions

Since studies employing SIR-type models are ubiquitous, it brings no cost to revisit how such a typical model physically interprets. Extended details will help readers walk through the model derivation, such that they are able to critically assess the assumptions and possible caveats.

### 2.1 Compartmentalization

We consider a region Ω that is endemic to COVID-19, occupied by humans of a constant total population *N*. The population is subdivided into several compartments based on the infection status and severity. The first compartment is *susceptible S*, determining a group of humans who are free from the virus but vulnerable to infections. An *S* –individual can have a first contact to the virus and it takes a few days from such an onset until symptoms appear, called *incubation period* θ^−1^ [d], during which the person belongs to the *exposed* compartment *E*. As COVID-19 also contracts asymptomatic cases (Rothe et al., 2020), at the termination of the incubation period, the person can still be asymptomatic *A* or symptomatic *I* depending on the immune response (fitness), age, and possible comorbidities (Huang et al., 2020). We assume that the asymptomatic cases share the proportion of *a* to the exposed cases. An asymptomatic case goes further to either of the two cases: *detected pA* or *undetected* (1 − *p*)*A*. Determination of the average probability measure *p* is heavily contingent on whether the person initiates a self-report to the medical department or not. Having been designated infection-positive, the asymptomatic person initiates self-quarantine at a certain rate. When the person undergoes a symptomatic infection, we assume that (s)he is automatically directed to a hospital. At this stage, all humans occupying *pA* and *I* are reported in the media as *active cases*. The moment when the infection ends, all the corresponding active cases become *closed cases* in which infection-free humans either recover or die, who are then compiled into *R* and *I* compartment, respectively.

### 2.2 Direct transmission

We assume that all the human individuals are homogenous, meaning that those with certain ages and social ranks are not treated differently than the others. The time scale (unit) is assumed to be 1 day [d]. The window of observation is considered short enough that the resultant of births and natural deaths is ignorable. Focusing on certain countries, we seek to simulate our model starting after global travel restriction, where migrations and imports in the population are negligible. Therefore, the total human population is considered constant. We assume that all the undetected individuals (1−*p*)*A* are free to move around like the susceptible humans, while those who undergo quarantine among *pA* are not. All the *I*–individuals clearly stay in hospitals. Infection happens when a susceptible person unrelated to medical department encounters an *A*–individual. A susceptible medical staff can also get infected through contact with an *I*–individual. Suppose that all the *A*– and *I*–individuals express certain sneezing behavior during infection with a uniform *sneezing rate s* [d^−1^]. This number determines how often they sneeze per day on average. Other important parameters for the successful virus transmission are an *effectivity measure e*_*s*_, determining how probable a sneeze (water droplets) leading to a successful infection, and a *probability measure p*_*s*_ determining how probable an infected person sneezes during contact with a susceptible person. Another parameter leading to successful infection is related to how proximate a susceptible and an infected person during contact are. We define a decreasing function *ϕ* (*r*) of the minimum contact radius *r* [m] describing how essential certain contact radii leading to the transfer of water droplets from an infected to a susceptible person. Moreover, we note that *A*/*N* determines the probability of a susceptible person to meet with an infected person from *A*, equalling to the number of *A*–individuals divided by the total population. The frequency multiplied by the probability, *S · A*/*N*, determines the ‘expected’ number of encounters that would probably lead to infection. This number, however, assumes that every *S* –individual encounters an *A*–individual only at maximum one time per day. If *k* denotes the average number of encounters with *A*–individuals experienced by any *S* –individual, then *k S A*/*N* denotes the average number of encounters that possibly cause infection. This seemingly large *k* (can also be greater than *N*/*A*) is neutralized by *p*_*s*_, and essentially the remaining parameters mentioned earlier, such that not all *S* –individuals experience infection per day. Therefore, attributed to direct transmission is the loss of individuals from the susceptible compartment *S* of daily magnitude

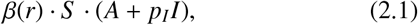

where *β*(*r*) := *s*· *e*_*s*_· *p*_*s*_· ϕ(*r*) · *k*· 1/*N*. One can think of the importance of *s, e*_*s*_, *p*_*s*_, ϕ(*r*) by asking if infection still happens when every of them is assumed vanishing. For example, if the sneezing rate *s* = 0, then no infection is prevailing since nobody sneezes during contact. Even when an infected person sneezes frequently, no infection can be expected if the sneezing is not done in front of a susceptible person, i.e., *p*_*s*_ = 0. The appearance of *p*_*I*_ in (2.1) stems from the preserved manner and medical restrictions imposed on *I*–individuals in hospitals.

### 2.3 Model for the infection rate

The new *infection rate β* clearly follows the behavior of ϕ with respect to the minimum contact radius *r*. Here, the function *β* is modeled in the sense of subtantiating certain statistical sampling. A sufficiently large number of contact cases of preferably different contact radii are pooled and the classified patients are distributed with respect to the minimum contact radius. We assume that the resulting distribution is superimposed by a normal distribution

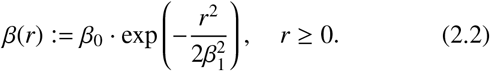

The parameter *β*_0_ defines a reference maximal infection rate obtained when the minimum contact radius *r* is 0. This parameter can be computed e.g. during an earlier take-off period where only a small number of humans are well-acknowledged with the aid of physical distancing. The other parameter *β*_1_ accounts for the switch from a significant decrement (*r* < *β*_1_ regime) to a slow decrement (*r* > *β*_1_ regime), indicating an interface between essential and non-essential physical distancing.

### 2.4 Fomite transmission

Experts hypothesize that the virus transmission is attributed to not only direct transmission but also *fomite transmission*. The latter represents transmission resulted from the act of touching the former-trace of sneezes from infected persons on surfaces. A recent study suggests that SARS-CoV-2 can survive on surfaces like plastic or stainless steel for up to *σ* ≈ 3d (van Doremalen et al., 2020) or, assuming similarity with SARS-CoV-1 and MERS-CoV, even as long as 9d (Kampf et al., 2020). Suppose that *A*–individuals in the past *σ*d walked around and sneezed where every individual created the average total sneezing coverage *c*_*σ*_ = *s*· *D*_*s*_ ·*c* _*f*_ [m^2^ ·d^−1^] per day, where *s* was the sneezing rate. The parameter *D*_*s*_ [m^2^] denotes the average unit area covered by a single sneeze. Samples for *D*_*s*_ can possibly be found on desks, chairs, beds, clothing, or door handles, cf. (CDC, 2020). The correction factor *c* _*f*_ clears out the possibility that the same individual or different individuals sneezed on the same spot. It thus is not expected that the accumulated area of the sneezes exceed the total area |Ω|. Due to restricted movement and isolation, we assume *c*_*σ*_ ≃ 0 corresponding to *I*–individuals in hospitals. With such composition, the following formula

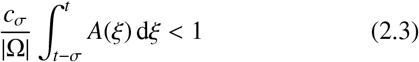

estimates the probability of a susceptible person in the current time *t* to encounter (not yet touch) the accumulated area for the former-trace of the sneezes from *A*–individuals from the past *σ*d. A sample upper bound for *c*_*σ*_ is gained from the fact that *A* ≤ *N* for all time, such that *c*_*σ*_*Nσ*/ |Ω| < 1 implies (2.3). The expression in (2.3) multiplied by the frequency *S*, the rate representing the daily intensity of touching surfaces *f*_*t*_, the probability of touching nose or mouth shortly after touching surfaces *p*_*nm*_, and the effectivity measure *e*_*s*_, returns the expected number of infection cases attributed to touching the former-trace of sneezes additional to the direct transmission. For brevity, we introduce a folder *α* := *f*_*t*_· *p*_*nm*_· *e*_*s*_· *c*_*σ*_/ |Ω|. Accordingly, the number of new infections in the susceptible compartment *S* undergoes the following correction

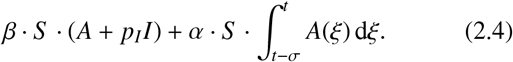

Covering unknowns, it is traditional to appoint the infection rates *β, α* for the fitting parameters.

### 2.5 Assumption on age structure

Age structure of COVID-19 dynamics abides by two mechanisms as variations of exposure to the virus and variations in clinical risk. Wu et al. (2020) investigate age-specific susceptibility to infections and fatality risk of symptomatic cases. Both measures showed a notable increase with age indicating highest portions for age-classes 60–69, 70–79 and > 79 years. Zhou et al. (2020) verify older age as a risk factor that leads to identifying vulnerable patients. They analyzed 191 patients in two hospitals in Wuhan to reveal that the median age was 56 years (IQR 46–67), while that of non-survivors and survivors were 69 (IQR 63–76) and 52 (IQR 45–58) respectively. These estimates also indicate higher exposure and fatality risk in higher ages. Verity et al. (2020) also report something similar in the case fatality rate. A considerable difference was evident in two age groups < 60 and ≥ 60 both from their parametric (1.4% and 4.5%) and non-parametric analysis (1.5% and 12.8%). This investigation was based on cases from both inside and outside of mainland China.

Knowing that the influence of age on the endemicity level cannot be disregarded – on the one hand – and the model can grow immensely in dimension due to age stratification – on the other hand – we infuse the information about age structure into the proportion of asymptomatic cases *a*. The preceding observations indicate that most symptomatic cases originate from older humans. The larger *a* means the more younger humans outside of hospitals spreading the virus.

### 2.6 Immunity

Researchers are still struggling to determine whether COVID-19 leaves out (partial) immunity to the recovered humans. The primary obstacle is as yet the acquired short course of the pandemic, as of time this paper is written. Research is still ongoing to see if COVID-19 is connected to SARS (2003) and MERS (2012) in terms of reaped immunity; if connections between both diseases extrapolate to certain behavior of COVID-19 is however not yet confirmed (Prompetchara et al., 2020). Considering the time obstacle, it thus is justifiable to assume that all recovered humans gain immunity during the relatively short observation in this study. As a (physical) consequence, there is no outflow from the recovered compartment directing to the susceptible compartment due to loss of immunity.

### 2.7 Household structure

We account for the difference in the virus transmission within and between households. The key idea is to break down all the state variables in the model into the household level. Let *i* denote the household index and *N*_*h*_ the average number of household members whose data can be accessed from the official website of the United Nations and that carrying census data in different countries (United Nations, 2019; Department of Census and Statistics - Sri Lanka, 2020). We assume there are *H* households occupying the observed region Ω. It is straightforward to see that *N* = *N*_*h*_*H*. Like in the population level, every individual can experience being susceptible, infected, recovered, or dead. We thus have *S* _*i*_, *E* _*i*_, *A* _*i*_, *I* _*i*_, *R* _*i*_, *D* _*i*_ for all *i* = 1,…, *H*. For brevity, we denote *S* := Σ_*i*_ *S*_*i*_, which also applies for the other state variables. We assume that close interactions within households define a larger number of encounters *k*_wh_ (wh stands for within-household) such that *k*_wh_ > *k*_ch_ (ch for cross-household). Consequently, the within-household *β*_*h*_ and cross-household infection rate *β* hold the relation *β*_*h*_ > *β* for all minimum contact radii *r*. For the sake of lowering the degree of unknowns in the model, we postulate

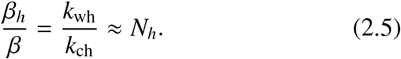

The approximation (2.5) describes certain behavioral overtones. We suppose that *k*_ch_ is relatively small and less sensitive to the change in the infection status due to an apparent border imposed on strangers rather than on household members. So, we fix *k*_ch_. When a standard household only contains *N*_*h*_ = 1 individual, then there is nobody in the household to encounter that would lead to new infection other than strangers. We then assume by (2.5) that the number of encounters per day doubles as now *N*_*h*_ = 2 individuals are present, triples as *N*_*h*_ = 3, and so on. An important remark is that this approximation may underestimate the real pictures as asymptomatic persons might still behave as if no infection occurs and if indeed so, the other members might take care of them incautiously.

As all the *I*–individuals stay in hospitals, it brings no relevance to cluster them based on household. However, they may still infect medical staffs. Decomposing the population on the household level gives us

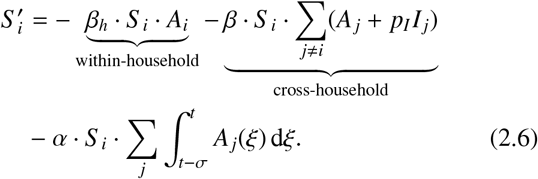

The third expression on the right-hand side of the equation is manifested from touching surfaces inside and outside home. Under a sufficiently large *H*, the sum in part of the second and in the third expression can nicely be folded into *p*_*I*_ *I* and 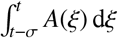, respectively. Violation to such an assumption might bear a certain outlier indexed by *i* where Σ _*j*≠*i*_ *p*_*I*_ *I* _*j*_ < *p*_*I*_ *I*. We further consider over the course of first three pandemic-months, that *A*_*i*_ ≪ *S* _*i*_ for almost all *i*. This assumption infers the situation where susceptible humans still dominate in most of the households. As a result, ∑_*i*_ *S* _*i*_*A*_*i*_≈ [(1/*H*) ∑_*i*_ *A*_*i*_]*S*. Making use of the estimate (2.5), we can sum up the expressions in (2.6) for all *i* to have

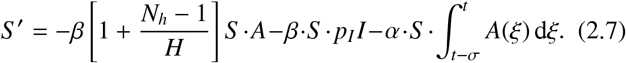

The effect of household structure can now be seen from the expression (*N*_*h*_ − 1)/*H*, which grows with *N*_*h*_. However, this expression turns to be negligible in the countrywide scale. Taking the case for Germany, we have (*N*_*h*_ − 1)/*H* ≈ 1/(4 × 10^7^). This negligible outcome was produced by violating our assumption that strangers with different health statuses would always have the possibility to encounter *k*_ch_ times a day on average, which is a serious defect. Practically, those from remote districts would have almost no chance to meet, yet they contribute in accelerations of the disease spread in their neighborhoods, which render a countrywide effect. Therefore, a correction is necessary to account for this issue. Let us denote Ω := ∪_*n*≤*C*_ Ω_*n*_ as a collection of different *C* localizations of the same number of households *H*_*C*_ and

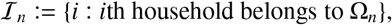

where *C* = *C*(*k*_ch_) and essentially (Ω_*n*_) have to be chosen such that the number of cross-household encounters *k*_ch_ is realistic. We acquire *H*_*C*_ = *H*/*C* and may also denote

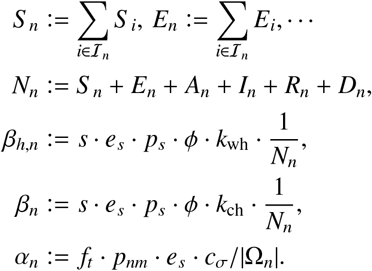

If we repeat the process as before, then we obtain a correction in (2.6) for *i* ∈ ℐ_*n*_ as follows

**Figure 2.1:**
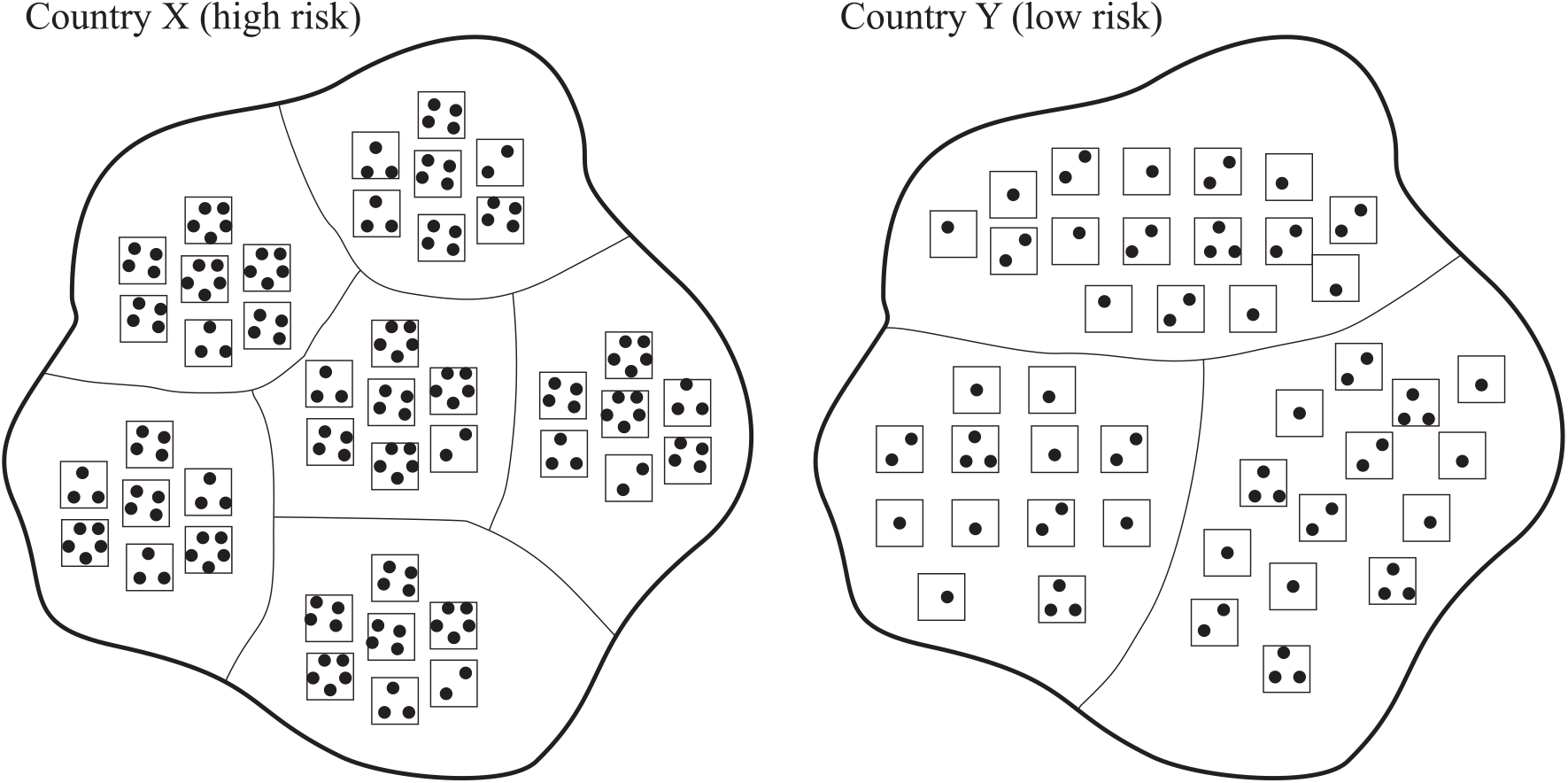
Two dummy countries of equivalent number of entire households, with different household structures and associated risk. Different localizations are bordered by thin lines; rectangles and small dots encode households and their members, respectively.

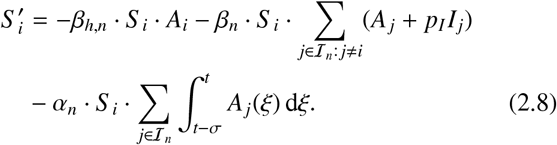

Under the dominance of susceptible humans, summing up the preceding equation over all *i* ∈ ℐ _*n*_ yields

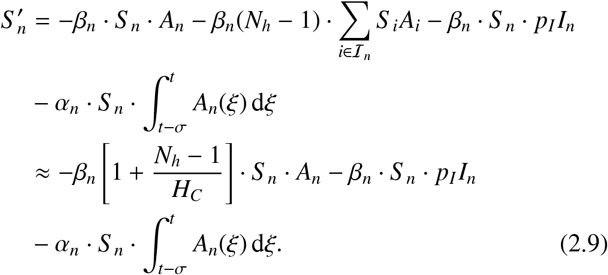

We can appoint *β, α* such that the summation of (2.9) over all *n* approximates

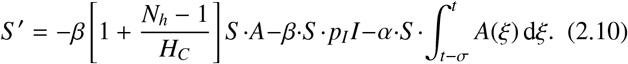

If *N*_*n*_ were constant for all *n*, then we have (*β, α*) = (*β*_*n*_, *α*_*n*_)/ *C*, which trace back our original infection rates. When *k*_ch_ is predetermined, surely can designating *C* as to represent blocks, subdistricts, districts, cities, provinces, or a whole country be handed over to decision-makers who know better to what extent most of the local inhabitants only interact with their own such that *k*_ch_ is realistic. Yet, this information might still be hard to get. From the point of view of data fitting, we can simply appoint the number of local households *H*_*C*_ as any constant.

Then, *k*_ch_ adjusts accordingly together with the other parameters as we seek to optimize the folder *β*. There is another aspect that we can extract from (2.10). Suppose that we compare two (dummy) countries X, Y of the same number of entire households *H*, see Fig. 2.1. Country X has citizens who have bigger families (large *N*_*h*_), like to stay around home, work nearby home, and are less dynamic in terms of meeting people other than staying home and working. This is the case where daily citizens’ spatial coverage is relatively low, inducing small |Ω_*n*_| ∼ 1/*C* ∼ *H*_*C*_. Country Y has more dynamic citizens of the same cross-household encounters *k*_ch_ (facilitated by adequate transportation system) as that of country X, also small families (small *N*_*h*_), work in more distant locations from home. The larger spatial coverage induces a large *H*_*C*_. In comparison, country X has a larger population as compared to country Y such that the same *k*_ch_ has to be acquired by country X and country Y with “less” and “much” effort, respectively. Our model (2.10) hypothesizes that country X poses higher risk of accelerated infection propagation due to its packed-type household structure and less dynamics as compared to country Y. The risk can also be understood in a different way. We can impose the same number of local households *H*_*C*_ but different *H*’s. In this setting, country X produces a larger *k*_ch_ than what country Y does. Prominent examples could be Sri Lanka–Germany or China–Finland. Looking at the same neighborhood of e.g. *H*_*C*_ = 100 households in those countries in comparison would justify the intuition on having different *k*_ch_’s.

### 2.8 Self-quarantine

All the humans who are detected to have been exposed to the virus proceed to quarantine for the period of *ν*^−1^ [d]. A study of 425 first patients in Wuhan, China has found that the mean incubation period from the distribution was 5.2d (Li et al., 2020a), but quarantine is suggested to take up 14d worldwide (Lauer et al., 2020), which is at the 99th percentile of the overall patients in the study. This means that they are obliged to stay at home and practice good hand during those days. Details such as keeping the body and hygiene-related personal belongings away from other members in the household as well as not having visitors should be apparent. Due to quarantine, as many as (1 − *ν*)*pA* detected infected individuals per day literally never go out for walk or even meet other household members and strangers. A wishful consequence is that now only *νpA* + (1 − *p*)*A* non-quarantined and undetected individuals are able to transmit the virus. Denoting *p*_*A*_ := *νp* + (1 − *p*), the infection term (2.10) changes to

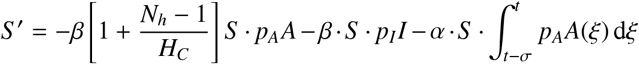

For a far-from-realistic case where the standard quarantine takes place ∞d, nobody from detected cases contributes to new infection via either direct or fomite transmission.

### 2.9 Model summary

Now our parameter *β* refers to the cross-household infection rate. To finalize the model, we assume no short movements from *A*– to *I*–individuals or vice versa during the illness periods and that mild health impacts on asymptomatic cases produce no deaths. All the preceding model descriptions translate to the following system

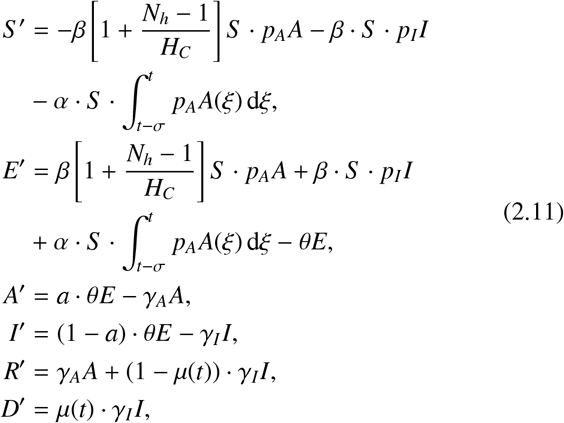

One can easily verify that every unit normal of the boundary of the nonnegative orthant in ℝ^6^ possesses a right-to-obtuse angle against the vector field of (2.11) in the corresponding boundary. This elucidates nonnegativity of the system trajectories for all possible parameter values, invoking a certain biological meaningfulness. The model also annexes practical relevance as the recovered *R* and death compartment *D* follow the cumulative mode owing to the fractional online case fatality rate *µ*(*t*). The initial conditions for all the state variables are either zero or positive depending on the initial time *t* = 0 taken.

### 2.10 Case fatality rate

Case fatality rate (CFR) usually is used to not only measure the deadliness of a certain infectious disease but also describe the general patients’ fitness as well as the tenacity of the health system in the observed region. A blunt connection to our modeling study is the fact that CFR dissevers the proportions of recovered and dead humans after average illness periods 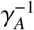 [d] for *A*–individuals and 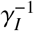 [d] for *I*–individuals. The actual CFR is to be calculated when the entire course of epidemics is completed, leaving out *only* closed cases (deaths and recoveries). Then, the value takes the ratio between total deaths and closed cases (Ghani et al., 2005). If the disease is still ongoing, computation of CFR becomes tricky. The widely-used estimate for its simplicity is called *naïve estimator*, calculated based on the ratio between accumulated deaths and confirmed cases over the entire history up to the current time. This ratio is biased and misleading, given that the outcome of part of the denominator is unknown, making the denominator grows much faster than the numerator (Angelopoulos et al., 2020). This fallacy has invited (statistical) researchers to come up with alternatives in the modification of the numerator and denominator, one claims advantages against the others. Speculations are based on whether concentrating on active cases and/or just closed cases (Ghani et al., 2005; Angelopoulos et al., 2020), age-dependence (Verity et al., 2020), time delay in between the real incidences and report (Baud et al., 2020), and different countrywise treatments in recording incidences (Johns Hopkins University, 2020b).

Our present study is not in the business of coming up with a correct, unbiased CFR. The fitting is heavily bound to data with no requirement for secondary analysis. Therefore, our task is to identify how the corresponding publisher reports the data in terms of estimating deaths and recoveries. In the model (2.11), the portion γ_*I*_ *I* who move out from hospitals per day are distributed to deaths and recoveries based on the online case fatality rate *µ*(*t*). Unlike usual CFR measures that relate to accumulated masses, this *µ*(*t*) corresponds to the division into deaths and recoveries only at the given time. These daily data can be found from the difference of the closed cases at the present and those at the previous time. If both accumulated deaths and recoveries do not advance after a day before, then we assign *µ* = 0 on that day. Finally, our prediction interprets how the data in the publisher would appear, not how the actual cases would behave.

### 2.11 Mass control

Governments worldwide have ordered their citizens to stay at home and cancel nonessential businesses to reduce the risk of infection. Curfew, lockdown, workplace clearance, and school closure are colloquial terms that converge to the same aim: avoiding mass gathering. Only then every term is uniquely determined by certain types of action. For example, lockdown is always associated with restricting people’s movement (avoiding going out from home), transportations, and business operations. In this study, we model these actions as to reduce the average number of cross-household encounters *k*_ch_. Added with law enforcement, we assume that people react in an impulsive way following the government’s order as per the commencement of either of those actions. The actions remain in effect unless they are closed by the government. We can then model *β* as a piecewise constant function over time, for a given minimum contact radius *r*. The constancy remains when there is no new action introduced in the population. Additionally, attention must be paid to the fact that the effect of a certain action can only be seen from incidence data given some time delay for people to get admitted to hospitals (such that they can be recorded) and for the recorded data to appear in the media. Such time delays can be fixed according to the incubation period.

## 3. The basic reproductive number

The following descriptions comprise details on the derivation of the basic reproductive number during an earlier take-off period. The key idea in calculating the basic reproductive number lies in finding a certain threshold at the interface of different stability statuses of the disease-free equilibrium (van den Driessche and Watmough, 2002).

Due to the constancy of the total population *N*, we can exclude *S* from the analysis by denoting *S* = *N* −*E* −*A* −*I* −*R* −*D*. We then seek to compute the basic reproductive number on an earlier take-off period, where one can assume that the online case fatality rate *µ* is relatively constant or simply *µ* = 0, and *R*(*t*) = *D*(*t*) = 0 for all − *σ* ≤ *t* ≤ 0. As a result, the subsystem (*R*^′^, *D*^′^) = (0, 0) degenerates into the equilibrium state (*R, D*) = 0. For an easy treatment, let us fold (*E*′, *A*′, *I*′)| _*R*=*D*=0_ into

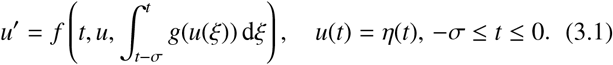

Substituting *E* = *A* = *I* = 0 into the right-hand side of (3.1), we obtain the disease-free equilibrium *u*_0_ = (0, 0, 0). The error 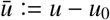 satisfies the equation

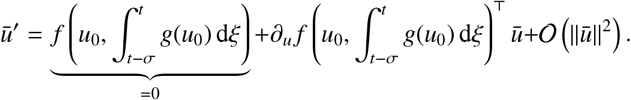

When a solution trajectory *u* is sufficiently close to *u*_0_, then the second-order term of the error is negligible and the Jacobian matrix

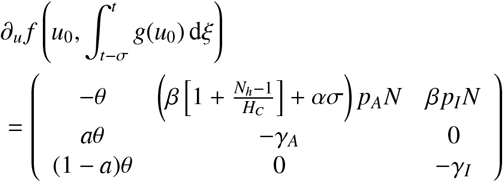

takes the lead in determining whether the trajectory will stay around *u*_0_ for all the remaining time. The trajectories can confine to *u*_0_ providing that all the eigenvalues of the Jacobian matrix have strictly negative real part. Apparently, the eigenvalues satisfy the following cubic equation

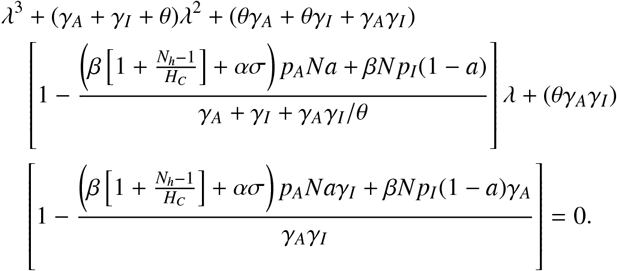

It is not difficult to show that all the coefficients of the cubic polynomial remain positive if the constant term is positive or the *basic reproductive number ℛ*_0_ < 1, where

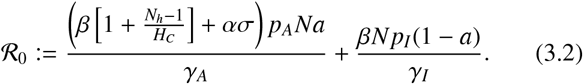

A direct consequence from having positive coefficients is that the polynomial intersects *y*–axis at a positive value and has positive first and second derivative for all positive λ, meaning there cannot be any root with positive real part. As the equilibrium state *u*_0_ is asymptotically stable in (3.1), the disease-free equilibrium (*E, A, I, R, D*) = (0, 0, 0, 0, 0) is also asymptotically stable in the coupled system (2.11), see (Vidyasagar, 1980, Theorem 3.1). The first term in the formula (3.2) represents a certain proportion 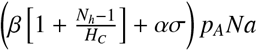 of new infection cases in the susceptible compartment attributed to an *A*–individual happening per an illness period 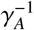. Observe that both direct and fomite transmission play role in increasing the proportion, which is enhanced as the virus would have survived longer (large *σ*) and there are more humans in the standard household (large *N*_*h*_). Surely is the transmission compromised as the proportion of asymptomatic cases is negligible (small *a*). The second term represents a similar contribution from an *I*– individual in a hospital, which of course is regulated by the probabilistic term *p*_*I*_ and their proportion (1−*a*).

Two important remarks are enumerated. First, the formulation of the basic reproductive number is restricted to the assumptions inherent from the earlier take-off period, i.e., *µ* = 0 and *R*(*t*) = *D*(*t*) = 0 for −*σ*≤ *t* ≤0. Therefore, the formula cannot afford its use on which the disease already advances through returning more deaths and intervention measures are overwhelming. Nonetheless, this early estimation is important for the understanding of the disease progression without any way of obstruction. How this value changes thus relies on the magnitude and tenacity of intervention measures taken in the next step. Second, the role of household structure is now more apparent. Two countries of the same choice of the number of local households *H*_*C*_ were predicted to exhibit different numbers of cross-household encounters *k*_ch_. When both high-risk and low-risk country are similar in other parameters except *k*_ch_ and *N*_*h*_ – which is almost the usual case – the values of *βN* from both countries should be significantly different: the high-risk country takes a larger value than the low-risk country. Amplified by *N*_*h*_, the basic reproductive number in the high-risk country becomes much larger than that in the low-risk country.

## 4. Study areas

### 4.1 Germany

The first COVID-19 confirmed case was reported on January 27, 2020 in Bavaria. The first two COVID-19-manifested deaths were reported on March 9, 2020 (Theresa Weiss, 2020). Entry restrictions and strict controls on borders were applied from the effect of March 15, 2020 and the German states closed all the schools, kindergartens, and universities on March 14, 2020 (Marcus, 2020). The Robert Koch Institute (RKI) alarmed the health risk level for residents in Germany from moderate to high on March 17, 2020 (Marcus, 2020). Bavaria and Saarland imposed curfew on March 21 (Marcus, 2020). Chancellor of Germany, Angela Merkel, announced a contact ban on March 22 late afternoon. On April 4, 200,000 stranded German vacationers return to Germany (Theresa Weiss, 2020).

### 4.2 Sri Lanka

First COVID-19 positive case reported in January 27, 2020 in Sri Lanka was a tourist from Hubei province, China. There were no reported cases until a first Sri Lankan was tested positive on March 11 (Epidemiology Unit - Sri Lanka, 2020). Then, the government took immediate actions to prevent further transmission by first closing the schools on March 12 followed by declaring public and mercantile holidays from March 16 and announcing work from home since March 20 (Department of Government Information - Sri Lanka, 2020). All passenger arrivals were banned on March 18 as all preliminary cases were reported among those returnees from Europe, Middle-East and India. All returnees have been sending to compulsory 2-week quarantine at designated centers operated by military under the instruction of health authorities. Those who completed quarantine has been further requested to undergo another 2-week self-quarantine too. Most critical restriction in public level was the island-wide curfew imposed from March 20 (Presidential Secretariat - Sri Lanka, 2020). Although the curfew has been lifting time-to-time, several districts including highly populated Western province have been under continuous curfew (Department of Government Information - Sri Lanka, 2020).

## 5. Numerical implementations

### 5.1 Problem definition

The data used in our investigation are collected by Johns Hopkins University (Dong et al., 2020), which can be accessed via a GitHub repository (GitHub, 2020). Three types of data are available: the accumulated number of deaths reported on a daily basis (`Dead`), accumulated recovered cases, and accumulated confirmed cases. The latter fold both accumulated deaths and recovered cases as well as active cases (`Active`) represented by *pA* + *I* in our model; see also further explanation in (Johns Hopkins University, 2020a). Observe that the model (2.11) already governs the timely accumulated deaths due to the absence of outflows from the compartment. Let us denote *t*_*s*_, *t* _*f*_ as the starting and final time for the assimilation, which are flexibly chosen according to one’s future need. Then, our aim is to find a set of parameter values ρ that solves the following problem

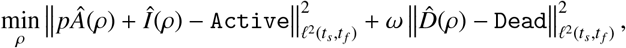

where *Â*’s, Î’s, and 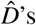‘s behavior are regulated by our model and ρ. The hat symbol, e.g. *Î*, represents the numerically approximated value of *I*–individuals from the model, picked on a daily basis. Note that in one time scale [d], the numerical approximate also comprises evaluations at different time points in between. The symbol ℓ^2^(*t*_*s*_, *t* _*f*_) denotes the usual least-square for discrete entities. The regularization parameter ω accounts for the trade-off between the fitting to active cases and that to deaths. Typically, ω is driven by a belief to what extent either of the two datasets is more reliable than the other. Balance is gained from training the two expressions in the objective function such that

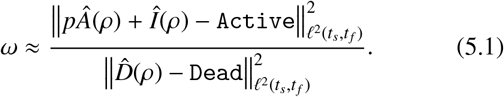

More details of the computation are provided in Sec. 5.3.

On the one hand, to what extent the infection rate *β* can make the model solution fits given data depends on some parameters, see (2.1). Covering unknowns, the infection rate has always been a good candidate for the fitting parameter (Lin et al., 2020). On the other hand, it still remains unclear if fomite transmission would also lead to new infection despite apparent indication of virus survival on surfaces (van Doremalen et al., 2020). Good empirical evidence was reported by Centers for Disease Control and Prevention (CDC). They confirmed that COVID-19 RNA was found on various surfaces in the Diamond Princess’s cabins of both symptomatic and asymptomatic cases even up to 17d posterior to clearance (CDC, 2020). Given this fact, human–virus contact tracing technically is an infeasible task, in which case the explanation continues with

> “… data cannot be used to determine whether transmission occurred from contaminated surfaces.”

We just see that the other infection rate *α* also deems unobservability. A relation between *β* and *α* is as yet beyond far from trivial.

The proportion of detected cases *p* for the case of China has been estimated around 14% (Li et al., 2020b). However, the value might vary from country to country due to different social awareness and mentality. Moreover, the proportion of asymptomatic cases *a* is naturally unobservable notwithstanding the previous assumption on age structure, i.e., that *a* contains information related to the ratio between young and older humans. The correction factor in the virus transmission between medical staffs and patients *p*_*I*_ is also unknown. Recall that 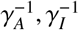 define the average illness periods spent by asymptomatic and symptomatic cases. All asymptomatic and hospitalized persons most likely leave their infection history uniquely, depending on their fitness, age, and available medical access. This information is both personal- and country-sensitive, in which case the average illness periods may also turn unobservable. In addition, the initial conditions for the exposed η_*E*_(*t*) and asymptomatic compartment η_*A*_(*t*) where *t*_*s*_ − *σ* ≤ *t* ≤ *t*_*s*_ are also unknown. We then chose the available data values in the past for the hospitalized (based on active cases, `Active` − *p*η_*A*_), recovered η_*R*_, and death compartment η_*D*_. The susceptible compartment clearly follows η_*S*_ = *N* − η_*E*_ − η_*A*_ – `Active` − *p*η_*A*_ − η_*R*_ − η_*D*_. The fitting parameters in this study are then summarized as

**Figure 4.1:**
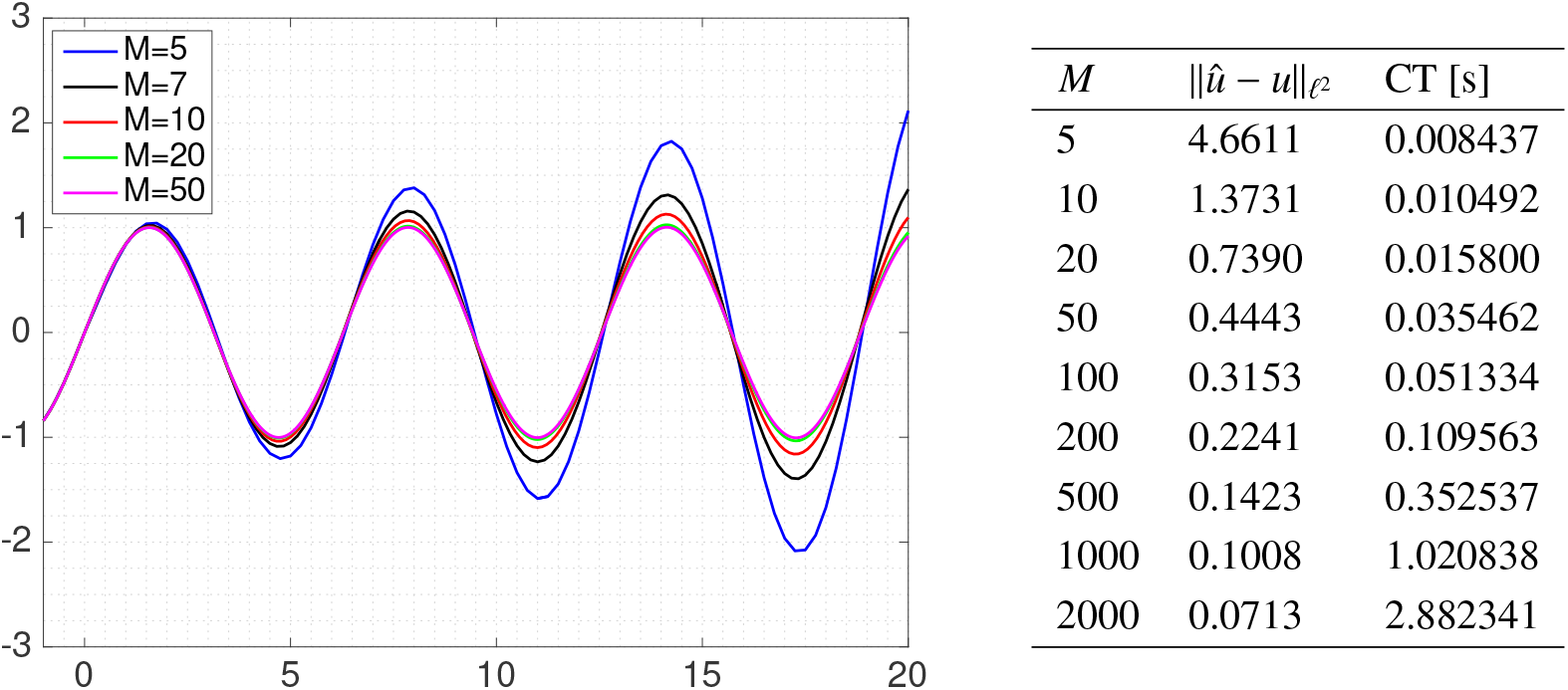
Numerical solutions of (5.2) using Pouzet-type SSPRK3 and different *M*’s calculated on a computer with the specification: Mac OSX 10.14, Processor 2.2GHz, RAM 8GB, Matlab 2015b. CT stands for computation time. A short observation indicates that the error accumulate as time grows.

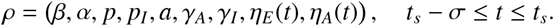

Finding optimal time-dependent parameters can be exhaustive since the number of actual parameters in the final discrete representation of the problem is dependent on the discretization taken. We avoid a possible curse of dimensionality by specifying parameter values on a daily basis, i.e., where historical data are defined, and determining those at time points in between using the spline interpolation.

### 5.2 Numerical treatment for the DIDE system

Our model system can be studied in the framework of a system of delay integro-differential equations (DIDEs) abstracted like in (3.1). We are in a good position to know that this type of system has been under previous investigations (Koto, 2002; Zhang and Vandewalle, 2004; Khasawneh and Mann, 2011; Shakourifar and Enright, 2011), even from the field of population dynamics (Kuang, 1993). A practical method for solving (3.1) is the direct adaptation of the (one-step) Runge–Kutta method, cf. (Koto, 2002). Without loss of generality, let us suppose that *t*_*s*_ = 0. We depart from the usual time discretization where the time domain for the initial condition [−*σ*, 0] is discretized into a set of, say, *M* grids of equal length. The time step and resulting time points of observation are then determined by *h* := *σ*/*M* and *t*_*n*_ := *hn, n* ∈ ℕ ∪ {0}. We assume equal length only for the sake of simplicity. The state variable *u* automatically undertakes the discretization *u* ≈ (*u*_0_, *u*_1_,…), containing six rows. A *d*–stage Runge–Kutta method features two vectors (*b*_*i*_), (*c*_*i*_) ∈ ℝ^*d*^ and one matrix (*a*_*i j*_) ∈ ℝ^*d*×*d*^ such that

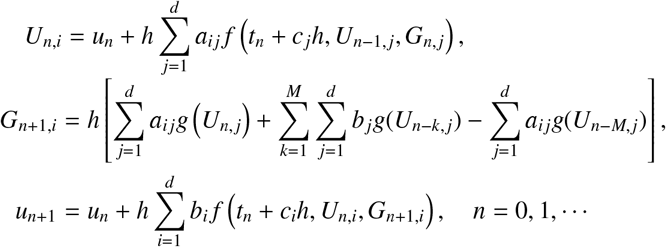

The initial conditions for the intermediate values *U*_*n,i*_ and approximate of the integral term *G*_0,*i*_ are given by

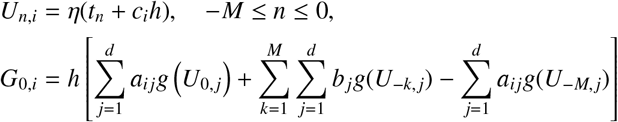

for all 1 ≤ *I* ≤ *d*. The scheme given above is of Pouzet-type where (*b*_*i*_), (*c*_*i*_), (*a*_*i j*_) are taken from the Runge–Kutta for ODE counterpart, and has been proven to be convergent (Brunner et al., 1982; Lubich, 1982).

Before applying the scheme to our model, we shortly test its performance. We consider a test function *u*(*t*) = sin(*t*) from which computations of the derivative and kernel term return the relation

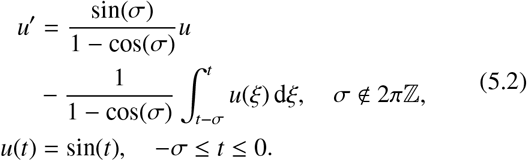

The time delay is specified as *σ* = 1 and starting and final time are *t*_*s*_ = 0, *t* _*f*_ = 20. The adopted scheme, also for the overall simulations in this paper, is Third-order Strong Stability Preserving Runge–Kutta (SSPRK3) represented by

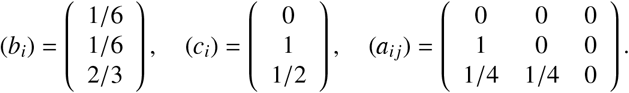

The computation results for different *M*’s are presented in Fig. 4.1. The numerical solution is named 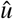.

### 5.3 Solving the parameter estimation problem

This study engineers a multi-parent genetic algorithm without the selection procedure (Ting, 2005). All the fitting parameters are represented via binary blocks (whose lengths depend on the boundary values), then concatenated to form a long binary chain representing a “player”. In every iteration, 50 players for ρ were used and the regularization parameter ω associated to the minimal objective function value is evaluated via (5.1). The mean from the whole iterations is then used to update the ω–value in the objective function, until then iterations are repeated. We then stop the iteration when the best objective function value gets stagnant after 25% of the total iterations that lead to the last constant value.

## 6. Numerical implementations

### 6.1 Fitting results

To accompany our modeling, we declare known parameter values in Tab. 5.1. The number of time grids in the domain for initial condition [−*σ*, 0] was chosen 300, meaning that every day we have 100 solution points with the starting point indicating the day. The time step used is then *h* = 0.01. However, for the comparison with data, we only assign the time points that correspond to the days. The number of local households was specified 100 for both Germany and Sri Lanka. Based on the average number of household members *N*_*h*_, Sri Lanka and Germany resemble the two dummy countries portrayed in Fig. 2.1. It is then expected that in a neighborhood of 100 households, Sri Lankans can easily encounter strangers/neighbours as compared to Germans or in particular, 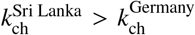. Unless irregularities are prevailing from the other parameters covered by *β*, we hypothesize that *β*^Sri Lanka^ > *β*^Germany^.

The unknown parameter values must be equipped with feasible ranges that are not too narrow (shorter binary chains or fast computation, but suboptimal) or too wide (slow computation, more optimal). We have designated for the earlier take-off period that 0 ≤ *β, α* ≤ 3/*N*, according to numerical experiments. Soon after a new action is introduced in the population, we assume that both *β* and *α* decrease reflecting people’s awareness and means of obeying such government’s order. All the probabilistic terms must satisfy 0 ≤ *p, p*_*I*_, *a* ≤ 1 for all time. According to (Guan et al., 2020), the median of hospitalization duration for the case of patients in China was 12d. For our numerical investigation, we impose 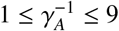 and 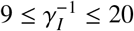 so the latter may vary around the cited 12d.

A phase is taken until there is a new countrywide action introduced in the population and its effect can be seen a few days after the commencement. We took 5 days for such a delay, which is approximately the incubation period. Therefore, we divide the time domain into four phases for Germany and three phases for Sri Lanka, each of which represents a subdomain where a certain action remains in effect, before added by another one. The fitting results for Germany and Sri Lanka can be seen in Figs. 5.1 and 5.4, respectively. Notwithstanding different numerical values yielded on every phase, our results support the hypothesis that *β*^Sri Lanka^ > *β*^Germany^ for all the overlapping time. The role of fomite transmission, which is steered by *α*, is much pronounced in Germany especially on earlier phases as compared to Sri Lanka. Our results allow us to argue that Germans could get infected through the comparable roles of direct and fomite transmission, whereas Sri Lankans are more overwhelmed by direct transmission through sneezing. The proportion of detected cases *p* for Germany (0.7976) is generally higher than that for Sri Lanka (0.5549). The correction factor related to the virus transmission from patients into medical staffs *p*_*I*_ generally starts from a small value during the earlier take-off period (0.0276 for Germany, 0.0048 for Sri Lanka). A hypothetical reason is due to a limited number of COVID-19 patients. Then, it waves to a significantly larger value (almost 1) and decreases thereafter. This means that, the within-hospital transmission entails almost no difference than the usual susceptible–asymptomatic transmission outside of hospitals in the recent phases. The mean values (0.3578 for Germany, 0.5790 for Sri Lanka) lead to the hypothesis that Sri Lankan medical staffs pose higher risk of getting infected in hospitals. A possible reason may be related to the reported irresponsible behavior of several COVID-19 patients who have been admitted to the hospitals without providing real information about their symptoms, historical whereabouts, and close connections to COVID-19 exposed individuals and even hiding from the health officials in backtracking (Arachchi, 2020a; Kariyawasam, 2020; Arachchi, 2020b). Moreover, the proportion of asymptomatic cases *a* is generally high for both countries (0.9778 for Germany, 0.9091 for Sri Lanka). This number weighs the exposed humans to be assigned to either the asymptomatic *A* or symptomatic *I* compartment in the optimal way such that the active cases *pA* + *I* match with data. This process is done without scrutinizing what happen to the *A* and *I* compartment individually. This modeling study owes correction to the value of *a* due to the unavailability of the hospitalized data for *I* with exact timing of admission and discharge. In case of presence, the objective function should be adjusted by including the hospitalized terms, and *a* should account for the tradeoff between active and hospitalized cases. Moreover, the findings on recovery rates lead to the hypothesis that the asymptomatic Germans suffer from illness for 5.14d on average, while the symptomatic ones for 18.94d. The asymptomatic and symptomatic Sri Lankans suffer from illness for 5.28d and 14.97d, respectively. Finally, the values of ℛ_0_ were computed only on the earlier take-off period. As mentioned in Sec. 3, the different household structures in Germany and Sri Lanka were shown to influence ℛ_0_ significantly. Our numerical results confirm this. Moreover, it should be noted that our ℛ_0_ was based on aggregating two proportions of new infection cases in the susceptible compartment from an undetected/non-quarantined *A*– individual and an *I*–individual, each per the corresponding illness period. Therefore, such a definition is not to be compared with the conventional geometric multiplier on the occurrence of infection cases per day attributed to a single infected person in a “virgin” population. For some more references on studies related to COVID-19’s ℛ_0_, see (Liu et al., 2020; Zhao et al., 2020; Adamik et al., 2020). We shall also note that ℛ_0_’s value is both model- and data-sensitive. At least, there has been no general proof that any distinct SIR-type models fitting equally well to the same data would produce identical values.

**Table 5.1:** Known parameter values from literature.

| Par | Description | Unit | Range | Germany | Sri Lanka | Reference |
| --- | --- | --- | --- | --- | --- | --- |
| $N$ | total population as of April 12, 2020 | [ind] | — | 83,726,366 | 21,395,610 | [worldometers.info](https://www.worldometers.info/) |
| $N_h$ | average number of household members | [ind] | — | 2.1 | 3.9 | (United Nations, 2019; Department of Census and Statistics - Sri Lanka, 2020) |
| $\nu^{-1}$ | quarantine period | [d] | 5.2–20 | 14 | 14 | (Lauer et al., 2020) |
| $\theta^{-1}$ | incubation period | [d] | 0–20 | 5.2 | 5.2 | (Li et al., 2020a) |
| $\sigma$ | duration of virus survival on surfaces | [d] | 0–9 | 3 | 3 | (van Doremalen et al., 2020) |
| $H_C$ | number of local households | — | — | 100 | 100 | |
| $M$ | number of time grids in $[-\sigma, 0]$ | — | $\geq 3$ | 300 | 300 | |

**Figure 5.1:**
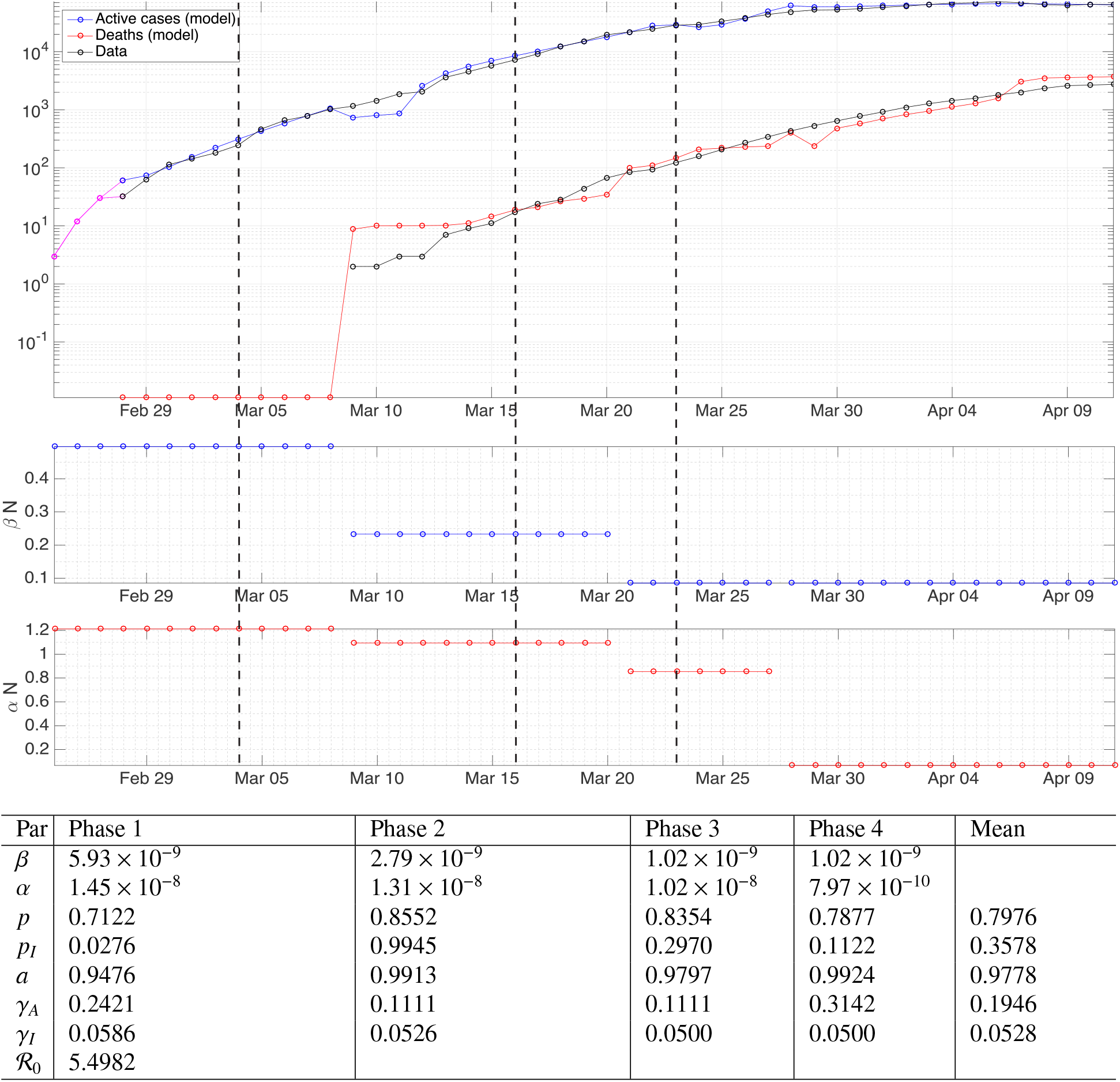
Fitting results using data for Germany. The magenta curve on the top panel shows the initial condition picked only at three points representing three days. Vertical dashed lines encode the time points where certain control measures are commenced: large events cancelled, school closure, and public curfew plus 1.5m minimum contact radius.

**Figure 5.2:**
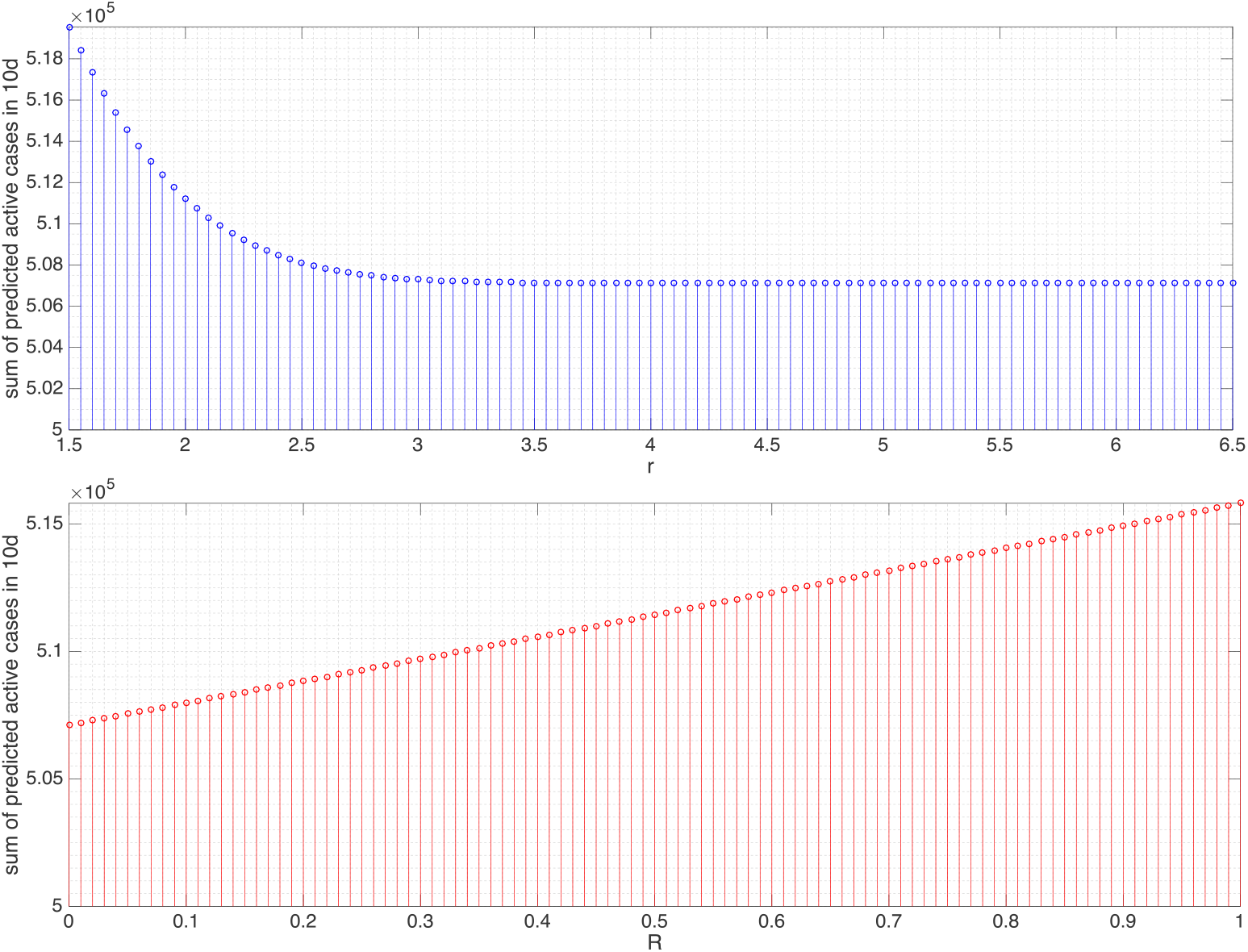
Sum of the predicted active cases for the next 10d in Germany under variations of *r* and *R*. The value *r* = 3 leads to the reduction to 98.31%, *R* ≈1/ 4, 1/3, 1/2, 3/4 to 98.73%, 98.86%, 99.15%, and 99.57%, respectively.

**Figure 5.3:**
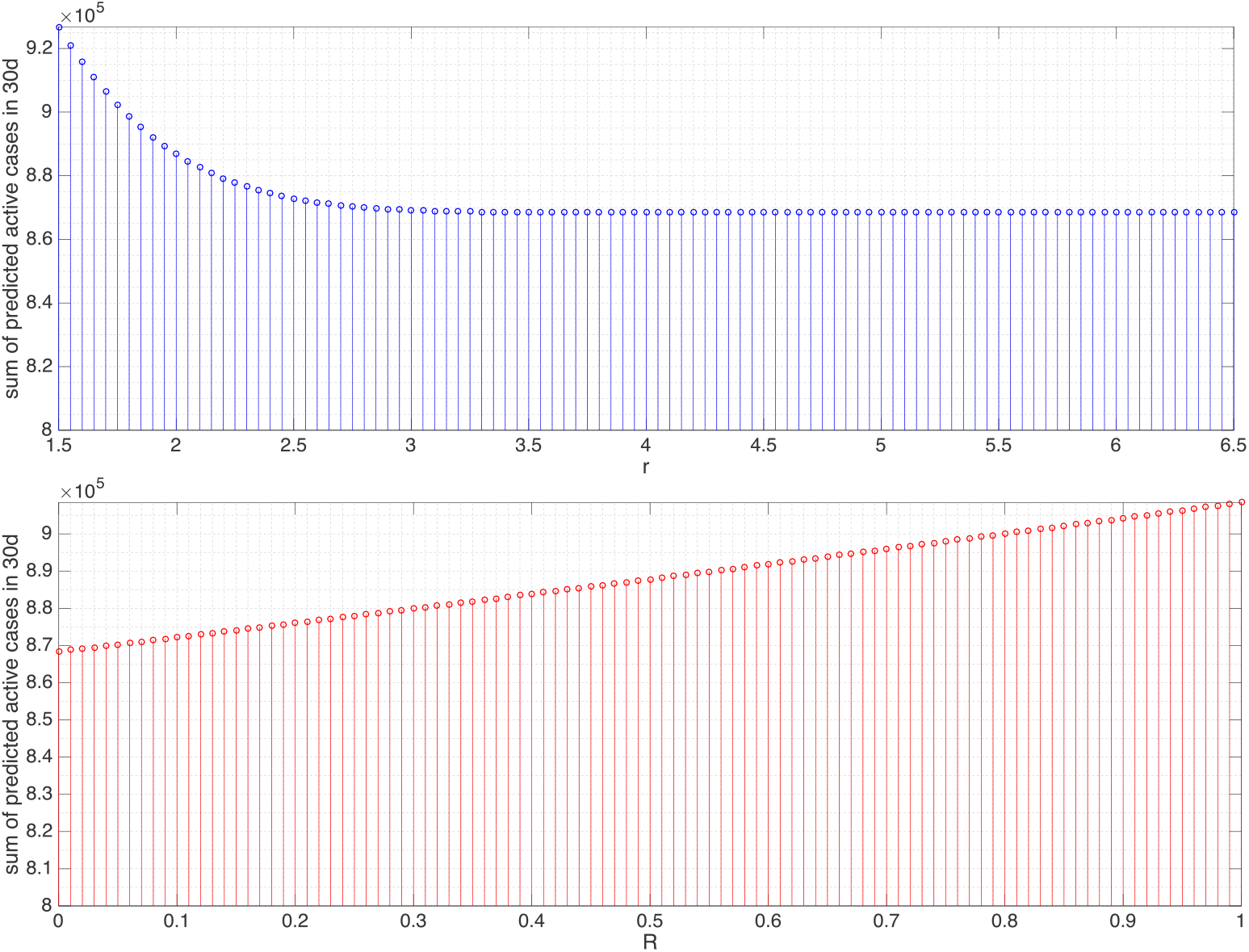
For the next 30d in Germany, *r* = 3 leads to the reduction to 95.59%, *R* ≈ 1/4, 1/3, 1/2, 3/4 to 96.65%, 96.99%, 97.73%, and 98.85%, respectively.

### 6.2. Readjustment of physical distancing and mass control

In Germany, public curfew and the application of 1.5m minimum contact radius were officially announced on March 22, 2020 late afternoon, commencing effectively the day after and remaining in effect five days later according to our hypothesis, which is the beginning of Phase 4. On the other hand, Phase 1 represents an earlier take-off period, where citizens were not bound to any official regulation regarding the physical distancing, inducing the minimum contact radius *r* = 0. We can now compute the deviation *β*_1_ in (2.2) by the identity 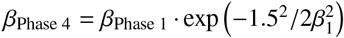, returning *β*_1_ ≈ 0.7997. This means that we are already in the inessential reduction of *β*. For Sri Lanka, official announcement regarding the social distancing was issued on March 20, 2020 (Presidential Secretariat - Sri Lanka, 2020), which lies in Phase 2. The minimum contact radius was assigned 1m in that announcement. We can then use a similar identity 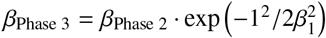 to reveal *β*_1_ ≈ 1.3299.

**Figure 5.4:**
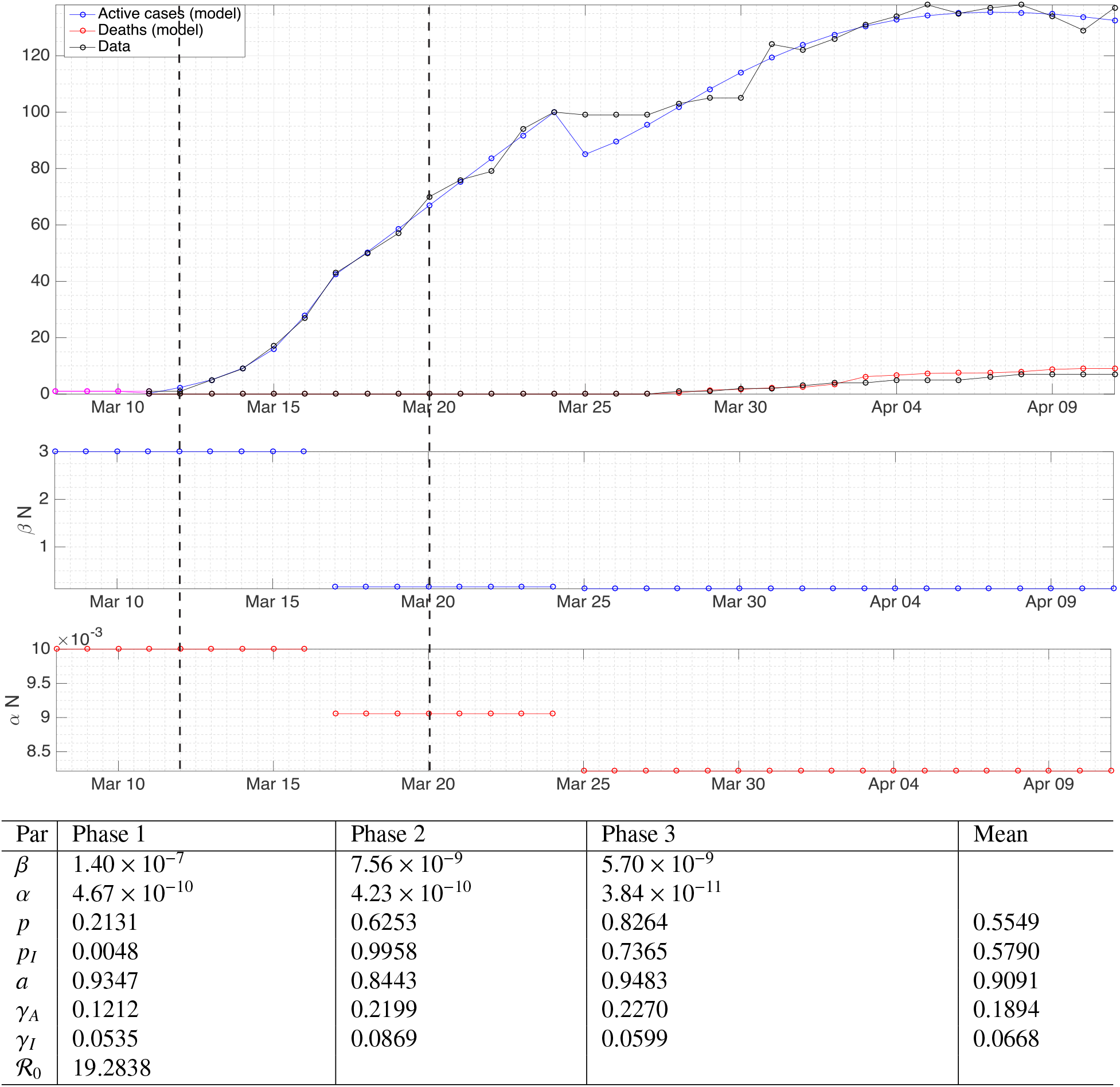
Fitting results using data for Sri Lanka. Vertical dashed lines encode the commencement of school closure and public curfew, respectively.

As the minimum contact radius is related to physical distancing, we seek to find a certain rule of thumb related to mass control (public curfew, lockdown, school closure, workplace clearance). As discussed in Sec. 2.11, the control can be implemented through reducing the cross-household encounters *k*_ch_. We introduce a ratio 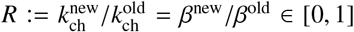 where *β*^old^ =*β*_phase 4_ for Germany and *β*^old^ =*β*_phase 3_ for Sri Lanka, also *R*=1 directs not to impose any reduction on *β*^old^. As *k*_ch_ denotes the ‘average’ number of cross-household encounters leading to possible infection, *R* can determine reduction on the intensity of all individual activities. For example, *R* = 1/ 2 means that every household member who goes for shopping twice a week must reduce to once a week, goes to office work six days per week must reduce to three days, does sport/outdoor exercise four times a week should change to twice a week, gets out from home twice a day changes to once a day, and so on.

The results from varying *r* and *R* in changing the sum of predicted active cases can be found in Figs. 5.2 and 5.3 for Germany and Figs. 5.5 and 5.6 for Sri Lanka. Several findings are highlighted. We did not attempt to increase *r* to be arbitrarily large as, apparently for both 10d- and 30d-prediction, the sums remain essentially constant after *r* = 3m for Germany and *r* = 4.5m for Sri Lanka. Our model hypothetically suggests that the minimum contact radius of 1.5m in Germany and 1m in Sri Lanka can still be improved to 3m and 4.5m, respectively, in order to gain significant reduction in the number of active cases. We understood that even though *β* can be reduced to zero by means of a lockdown, the residues in the portions of exposed and asymptomatic as well as symptomatic compartment eliminate exponentially but hardly jump to zero in an impulsive manner. This is why we can only afford reduction to the least of 95.59% for Germany and 78.90% for Sri Lanka. In Germany, increasing the minimum contact radius to 3m is equivalent in the outcome of predicted active cases to giving the intensity reduction ratio 1/4. This means that, every person who cannot take up 3m minimum contact radius can reduce his/her intensity of daily/weekly activities to 1/4 or vice versa. In Sri Lanka, predictions of the active cases in the next 10d and 30d give significantly different outcomes. For a short term (10d), extending the minimum contact radius to 4.5m gives more significant reduction (by 78.90%) as compared to giving the intensity reduction ratio 1/4 (93%). For a longer term (30d), both actions are equivalent in terms of the outcomes.

**Figure 5.5.**
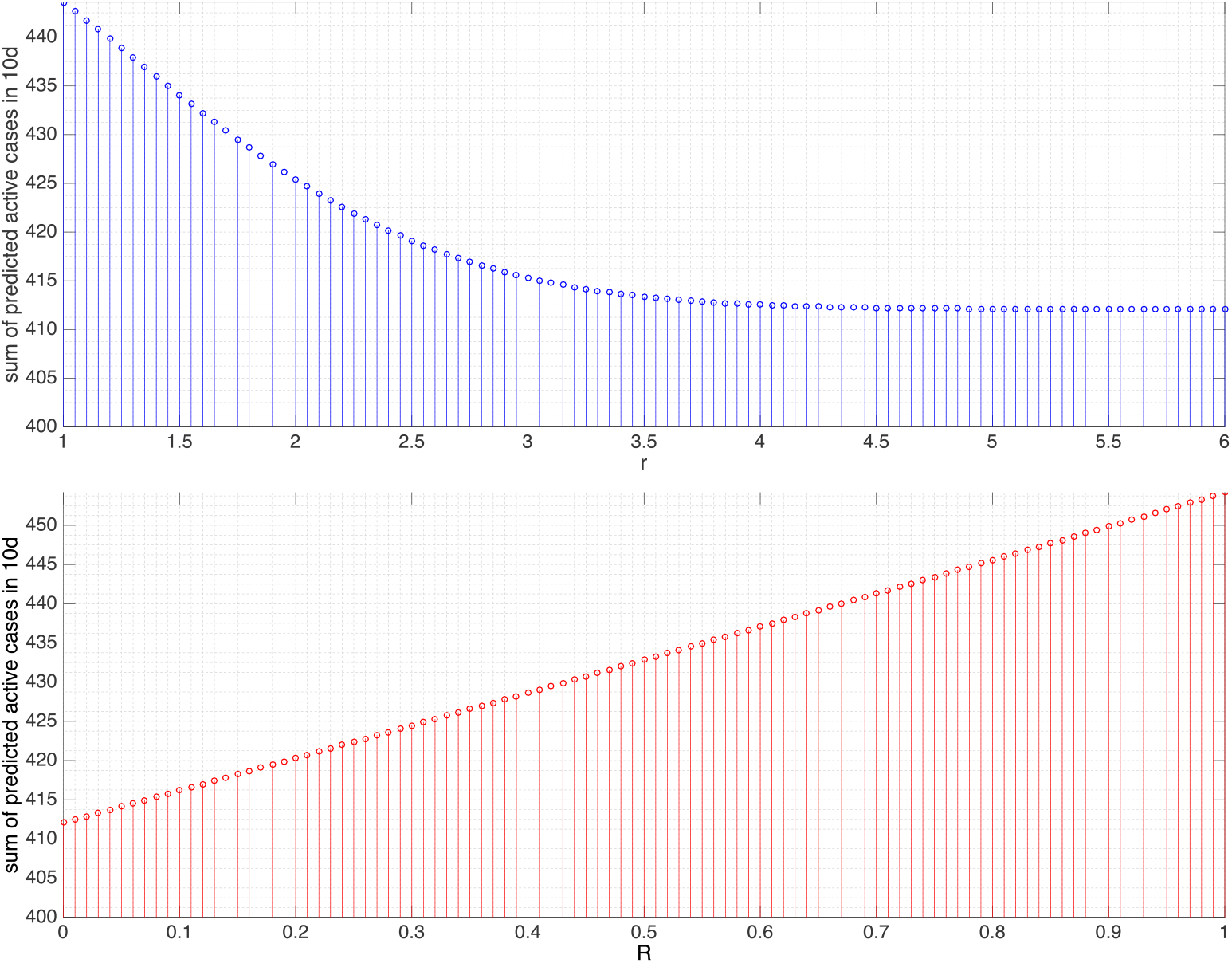
Sum of the predicted active cases for the next 10d in Sri Lanka under variation of *r* and *R*. The value *r* = 4.5 leads to the reduction to 78.90%, *R* ≈ 1/4, 1/3, 1/2, 3/4 to 93%, 93.73%,95.3%, and 97.63%, respectively.

**Figure 5.6.**
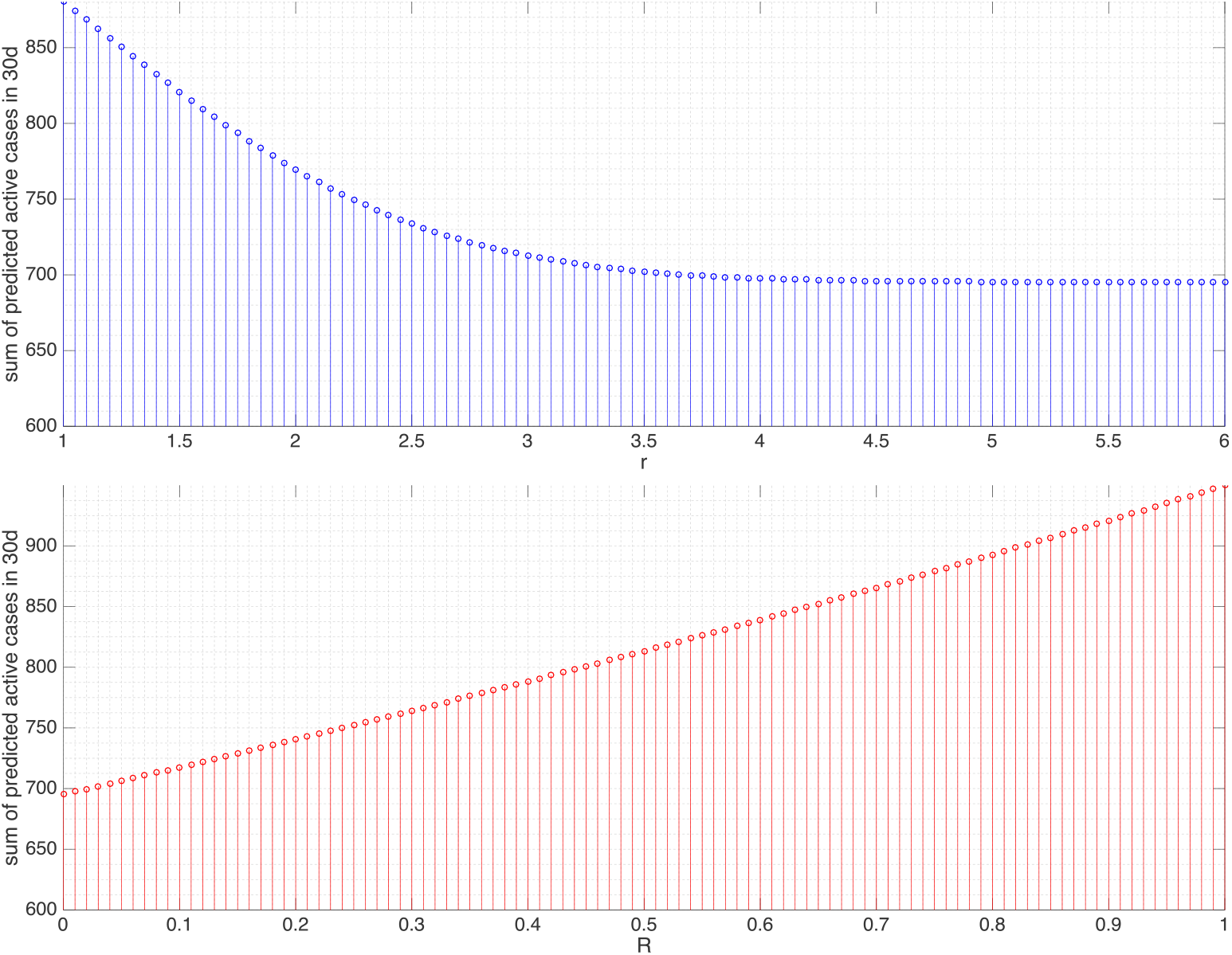
For the next 30d in Sri Lanka, *r* = 4.5 leads to the reduction to 78.90%, *R* ≈ 1/4, 1/3, 1/2, 3/4 to 79.19%, 81.2%, 85.64%, and 92.56%, respectively.

## 7. Conclusion

A micro-level approach on individual behavior and household structure in viewing COVID-19 transmission is proposed in this work. It caters linking infected status in more realistic way before compromising into macro-level. For instance, sneezing and surface touching are treated in a detailed manner. Meanwhile, we design the infection rate *β* in a unique way by amalgamating many factors relevant to direct transmission through sneezing. Manipulation of a decreasing function *β* of the minimum contact radius *r* bridges the scientific argument in propounding the physical distancing. Additionally, several notable factors are taken in modeling fomite transmission through surface touching. We enumerate the final effect of these as an accumulation over a period *σ*d in the past that asymptomatic individuals walked around and freely sneezed. Moreover, with the aim of tolerating differences to and fro households, we introduce the effect of different household structures into the model involving the number of local households and the average number of household members. This leads to easier identification of low-risk and high-risk country in terms of the basic reproductive number on an earlier take-off period.

Our model solutions using the final attribution (2.6) are in quite good agreement with available data from Germany and Sri Lanka. Unobservable effects that may represent reality, related to the optimal numerical values, have been hypothesized. The rule of thumb which we propose from our findings is that, considering long-term predictions over the active cases, Germany and Sri Lanka can ameliorate the endemicity level by increasing 1.5m to 3m and 1m to 4.5m minimum contact radius, respectively, and reducing the intensity of individual activities to 1/ 4 by any means of mass control. The latter should be the aspect the governments know better how to put into realization. Both measures can also be applied in combination flexibly, in the sense that those who cannot stick to one can apply the other. For some alternating choices between actions and outcomes, reduced magnitudes of the intensity of individual activities to 1/ 3, 1/2, 3/4 are also presented. Besides these choices, translating other intervention measures such as wearing mask in more secure way into equal reduction on *β* without having to stay away from friends/colleagues and minimize outdoor activities could be a possible outlook. Intensive cleaning of surfaces is also an additional intervention to mitigate fomite transmission.

Finally, we realized that the data used in this study as well as corresponding results will not infer a longer period ahead. In the sense that data jump to larger values after April 11 than predicted, our findings should remain actual and the actions might even need emphasis. When the data devolve to lower values, we have been let to observe based on the historical progression of the active cases that the jump may not be so radical as well. It thus puts no harm to keep the results as guidance, which may be subject to minor-to-medium infringement.

## Data Availability

Not applicable

